# What predicts adherence to COVID-19 government guidelines? Longitudinal analyses of 51,000 UK adults

**DOI:** 10.1101/2020.10.19.20215376

**Authors:** Liam Wright, Andrew Steptoe, Daisy Fancourt

**Affiliations:** Department of Behavioural Science and Health, University College London; Department of Epidemiology and Public Health, University College London

## Abstract

In the absence of a vaccine, governments have focused on social distancing, self-isolation, and increased hygiene procedures to reduce the transmission of SARS-CoV-2 (COVID-19). Compliance with these measures requires voluntary cooperation from citizens. Yet, compliance is not complete, and existing studies provide limited understanding of what factors influence compliance; in particular modifiable factors. We use weekly panel data from 51,000 adults across the first three months of lockdown in the UK to identify factors that are related to compliance with COVID-19 guidelines. We find evidence that increased confidence in government to tackle the pandemic is longitudinally related to higher compliance, but little evidence that factors such as mental health and wellbeing, worries about future adversities, and social isolation and loneliness are related to changes in compliance. Our results suggest that to effectively manage the pandemic, governments should ensure that confidence is maintained, something which has not occurred in all countries.

---

The pandemic of SARS-CoV-2 (COVID-19) has had wide ranging impacts on societies across the globe. In the absence of a vaccine, governments have implemented a range of measures to tackle the pandemic, generally focused on reducing transmission of the virus through: isolating those with diagnosed or suspected COVID-19, increasing ‘social distancing’ (e.g. working from home, restricting non-essential travel and limiting groups gathering in public venues), and enhancing hygiene procedures (such as the wearing of face masks). Existing evidence suggest the measures could have large impacts on infection rates and, subsequently, on reducing overall mortality ^1,2^. However, each of these measures requires citizens to make changes to their usual behaviour, sometimes at considerable personal cost ^3^. Though some measures have the force of law, in democratic societies unwilling to exercise authoritarian power, compliance requires voluntary cooperation. Yet, ensuring high levels of compliance has been a challenge ^4–6^. To manage the pandemic effectively, it is vital that we understand the factors that drive compliance; especially those factors that could be modifiable. There is a moderate literature from previous epidemics, such as the 2009 H1N1 swine flu and the 2003 severe acute respiratory syndrome (SARS), on factors that influence compliance with social distancing, hygiene, and quarantine rules ^7,8^. Several factors are highlighted in this literature, including trust and confidence in institutions, social experiences, mental health and wellbeing, and knowledge of the virus, each discussed below.

With regards to *confidence in institutions*, researchers research suggests that trust and confidence in government can increase compliance by assuring citizens that guidelines are necessary and effective ^9^. During the Ebola epidemic in Liberia, individuals with greater trust in government were more likely to comply with mandated social distancing ^10^, whilst during COVID-19, multiple cross-sectional studies have shown a small association between trust in governmental figures and compliance with protective measures ^11–13^. Reductions in geographical mobility – an indicator of adherence with lockdown and social distancing regulations – were also greater in regions of Europe with higher pre-pandemic levels of trust in politicians ^14^. However, a limitation of these studies is their focus on the early stages of the pandemic. A relationship was found between trust and compliance only at the start of the H1N1 pandemic in the Netherlands ^15^. Moreover, confidence may be a “double-edged sword” if citizens feel that success is assured regardless of their own actions ^16^.

Compliance may additionally be related to *mental health and wellbeing*, although empirical results and influential models of the relationship between positive affect and behaviour make conflicting predictions ^17^. On the one hand, depression and anxiety are related to lower extraversion, sociability ^18^ and increased risk aversion ^19,20^, which could increase compliance. On the other hand, depression is associated with lower self-efficacy and, in one study, lower altruism ^21,22^, factors which are associated with lower adherence, and depression is linked to non-compliance with medical treatments, more generally ^23^. Evidence from previous pandemics found that state anxiety was related to higher compliance ^7^, including a study from the SARS outbreak in Hong Kong using longitudinal assessment ^24^. Studies using cross-sectional data from Japan, China and the UK during the COVID-19 pandemic are less consistent ^25–27^, but one longitudinal study found an association between current life satisfaction and higher compliance ^17^, though lagged life satisfaction had the opposite association. The cause of poor mental health may also be relevant, with COVID-19-specific fears and worries (such as catching the virus) shown to be related to higher compliance ^11,25,28^, but other stressors, such as worrying about loss of employment, potentially leading individuals to break social distancing rules. Further, the relationship between mental health, wellbeing and compliance is likely bidirectional, as compliance with quarantine and social distancing may itself be a cause of poor mental health ^29^.

As another factor, the Health Belief Model ^30^ posits that health behaviour decisions are determined by weighing the costs and benefits of different courses of action against beliefs about one’s ability to enact these actions (i.e. self-efficacy). So *greater knowledge and exposure to information about COVID-19* may increase compliance by increasing awareness of the risks of catching COVID-19. However, it may also widen exposure to violations of social norms, which may discourage pro-social motivations to comply with guidelines ^31^, and to information that could induce fatalism ^32^. Several cross-sectional studies have already shown that knowledge about COVID-19 is related to greater self-reported compliance with preventative measures ^33–36^. However, these associations could be explained by individuals with greater willingness to comply also being more likely to seek out information about COVID-19. Notably, there is experimental evidence that providing higher expert estimates of infectiousness can *reduce* reported willingness to follow social distancing measures ^32^, suggesting that whether improved knowledge increases compliance depends on pre-existing beliefs. Individuals who seek out information may also be more trusting in general. Further cross-sectional and longitudinal studies show that beliefs about the efficacy of COVID-19 measures are associated with higher compliance ^12,13^.

A fourth group of factors that may affect compliance are *social experiences*. An important question is the extent to which social isolation impacts compliance, given that compliance with social distancing measures entail impacts on loneliness and on interpersonal contact ^29,37^. Loneliness is related to poorer health-practices and health-promoting behaviours, with behavioural disengagement in the face of stressors, and with lower perceptions of control ^38^. However, direct empirical evidence from the COVID-19 pandemic is limited. Loneliness has been shown to be cross-sectionally related to lower compliance ^39^, but given the possibility of reverse causality, longitudinal methods are required.

Finally, there is reason to believe that *behaviours and time-use* during quarantine could be related to compliance, likely in a bi-directional way. Some activities may make compliance more pleasant, reducing boredom, improving wellbeing, and incentivizing people to stay at home ^40^. Performing specific activities, such as working outside the home or caring for friends or relatives, may also pose a challenge for compliance, either due to the inevitable consequence of individuals’ careers (such as essential workers being unable to stay at home during strict lockdowns) or due to economic necessity. During the COVID-19 pandemic, studies have shown that reductions in population mobility were lower in poorer areas of the United States ^41^, and that compliance could be increased if individuals are financially compensated (Bodas and Peleg, 2020). Therefore, investigating how compliance co-varies with time use could help to identify groups who need more support to comply with guidelines.

Overall, then, there is preliminary evidence to suggest that a range of factors could be related to compliance during COVID-19. However, to date, most of the literature on compliance during COVID-19 has used cross-sectional data, which raises the possibility that associations are explained by reverse causality or unobserved confounding. Further, there has been little research directly comparing the size of association between different predictive factors and compliance. Such research is important for public health professionals and policy makers deciding what sorts of messaging and interventions are needed to maintain high adherence to try and control the spread of the virus. Therefore, in this paper we use data from a weekly panel of 51,600 adults across twelve weeks of lockdown in the UK (01 April – 22 June) to explore which factors out of a wide range drawn from the literature cited above were associated with self-reported adherence to government guidelines to tackle COVID-19. Specifically, we assessed the role of mental health and wellbeing, confidence in government and the health service, social isolation, stressors, such as fear of job loss and threats to personal safety, and knowledge of COVID-19. Our study presents a substantial advance on previous research by exploiting the longitudinal structure of our data to test for reverse causality and to account for time-invariant heterogeneity across individuals, and by using Random Intercept Cross-Lagged Panel Models (RI-CLPM)^42^ to assess whether *within-person* changes in potential predictors of compliance are related to later changes in compliance, a question that is more consistent with a causal process ^43^.

## Results

### Descriptive Statistics

Descriptive statistics on the sample characteristics, including age, gender, and income group, are displayed in Table 1 and further descriptive data on measures in the study are shown in Supplementary Table S1. Participants completed 8.9 interviews, on average. 38.62% of the sample had 11 or more interviews over the analysis period.

**Table 1:** Sample characteristics

|  |  | Unweighted |  | Weighted |  |
| --- | --- | --- | --- | --- | --- |
|  | Variable | Individuals (%) | Observations (%) | Individuals (%) | Observations (%) |
| Gender | Male | 12,737<br>(24.68%) | 95,617<br>(25.14%) | 24,915.95<br>(48.29%) | 186,780.10<br>(49.1%) |
|  | Female | 38,863<br>(75.32%) | 284,754<br>(74.86%) | 26,684.05<br>(51.71%) | 193,590.90<br>(50.9%) |
| Age group | 18-29 | 4,081<br>(7.91%) | 24,329<br>(6.4%) | 7,184.89<br>(13.92%) | 41,212.23<br>(10.83%) |
|  | 30-45 | 14,390<br>(27.89%) | 95,262<br>(25.04%) | 12,039.87<br>(23.33%) | 78,546.30<br>(20.65%) |
|  | 46-59 | 16,596<br>(32.16%) | 122,414<br>(32.18%) | 13,620.67<br>(26.4%) | 100,771.58<br>(26.49%) |
|  | 60+ | 16,533<br>(32.04%) | 138,366<br>(36.38%) | 18,754.57<br>(36.35%) | 159,840.89<br>(42.02%) |
| Highest qualification | GCSE or below | 7,005<br>(13.58%) | 51,298<br>(13.49%) | 16,086.29<br>(31.17%) | 119,670.27<br>(31.46%) |
|  | A-levels or equivalent | 9,019<br>(17.48%) | 65,311<br>(17.17%) | 17,029.29<br>(33%) | 122,955.73<br>(32.33%) |
|  | Degree or above | 35,576<br>(68.95%) | 263,762<br>(69.34%) | 18,484.42<br>(35.82%) | 137,744.99<br>(36.21%) |
| Ethnic group | White | 49,169<br>(95.29%) | 364,742<br>(95.89%) | 46,466.04<br>(90.05%) | 347,623.10<br>(91.39%) |
|  | Non-White | 2,431<br>(4.71%) | 15,629<br>(4.11%) | 5,133.96<br>(9.95%) | 32,747.90<br>(8.61%) |
| Household income | <£16k | 6,711<br>(13.01%) | 49,428<br>(12.99%) | 9,209.31<br>(17.85%) | 66,923.37<br>(17.59%) |
|  | £16k - £30k | 11,198<br>(21.7%) | 84,716<br>(22.27%) | 12,682.74<br>(24.58%) | 97,202.01<br>(25.55%) |

| Variable | Unweighted |  | Weighted |  |
| --- | --- | --- | --- | --- |
|  | Individuals (%) | Observations (%) | Individuals (%) | Observations (%) |
| £30k - £60k | 16,459<br>(31.9%) | 121,186<br>(31.86%) | 15,043.87<br>(29.15%) | 110,408.74<br>(29.03%) |
| £60k -£90k | 7,353<br>(14.25%) | 52,428<br>(13.78%) | 5,649.25<br>(10.95%) | 40,075.85<br>(10.54%) |
| £90+ | 5,131<br>(9.94%) | 36,600<br>(9.62%) | 3,657.11<br>(7.09%) | 25,800.31<br>(6.78%) |
| <NA> | 4,748<br>(9.2%) | 36,013<br>(9.47%) | 5,357.72<br>(10.38%) | 39,960.72<br>(10.51%) |

*Compliance with guidelines* was measured weekly using a single-item measure: “Are you following the recommendations from authorities to prevent spread of Covid-19?”, measured continuously from 1 = Not at all – 7 = Very much so. Details on the other measures in the analyses are presented in the Methods section.

Fit statistics for the RI-CLPM models are displayed in Table. While RMSEA statistics are lower than conventional cut-offs, SRMR, CFI and TLI scores are poor in most cases. This is likely to be partly due to the non-normally distributed variables used in this analysis. The average autoregressive effect of compliance upon later compliance in the RI-CLPM models was 0.34, indicating that one third of the change in compliance persisted to the next interview and suggesting cross-lagged effects did not dissipate immediately.

### Confidence in Institutions

When exploring cross-sectional “between” variation in compliance and confidence in institutions, higher confidence in government, the health service and access to essentials were all associated with higher compliance, although associations were small (right panel, Figure 2). However, when exploring the cross-lagged associations, an increase in confidence in government was related to a small 0.02 SD increase in compliance at the next interview, but no other factors were significantly related to compliance in later waves (left panel). Compliance did not predict changes in confidence in institutions (middle panel).

**Figure 1:**
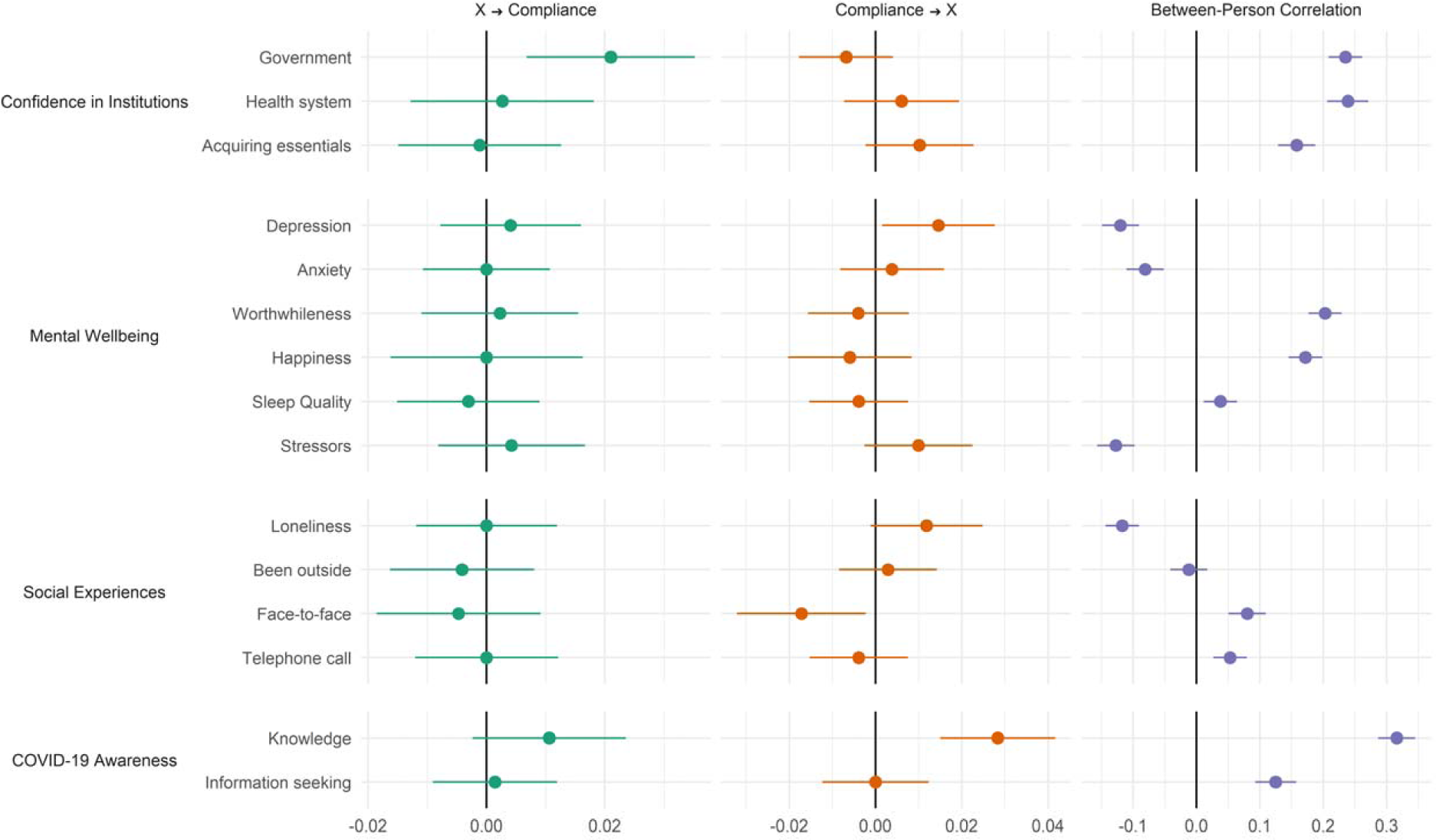
RI-CLPM Model Results. Left panel shows the cross-lagged effect of the exposure variable on compliance; the middle panel shows the cross-lagged effect of compliance on the exposure variable; and the right panel shows the correlation between the random intercept terms for the exposure and compliance (i.e. the between person correlations). Estimates are standardized.

**Figure 2:**
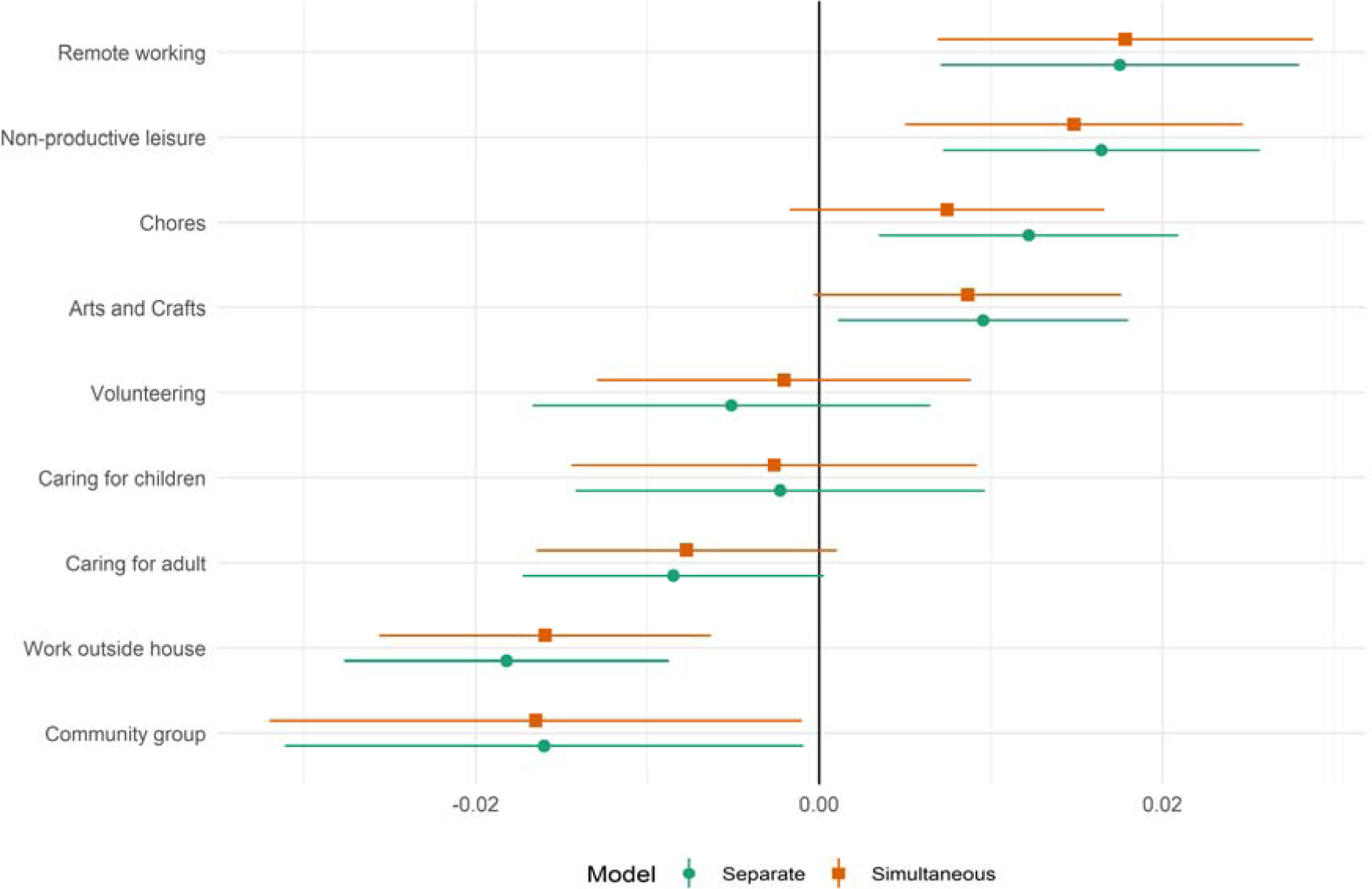
Standardized fixed effects model results, regression of compliance on time-use variables, entered separately or simultaneously into models.

### Mental wellbeing

Higher compliance was cross-sectionally related to lower depression, anxiety, and stressors, and higher happiness, sense that life’s activities were worthwhile, and sleep quality (Figure 2). However, when exploring the cross-lagged associations, no factors relating to mental wellbeing were associated with later improvements in compliance, and instead compliance was related to increases in depression.

### Social experiences

Higher compliance was cross-sectionally related to lower loneliness and higher face-to-face and telephone contact but not to time spent outdoors (Figure 2). However, when exploring the cross-lagged associations, no factors relating to social experiences were associated with later improvements in compliance, and instead compliance was related to decreases in face-to-face contact.

### COVID-19 awareness

Higher compliance was cross-sectionally related to higher levels of knowledge and information seeking relating to COVID-19 (Figure 2). However, when exploring the cross-lagged associations, no factors relating to COVID-19 awareness were associated with later improvements in compliance, and instead compliance was related to increases in knowledge about COVID-19.

### Time use

We assessed concurrent changes between time use and compliance levels using fixed effect models. estimates from these models are displayed in Figure 2. Working outside the house, caring for a friend or relative, and engaging with a community group were each associated with lower compliance, while time spent in arts and crafts, non-productive leisure, doing household chores, and working remotely were associated with higher compliance. Associations were generally little impacted by whether time-use variables were added separately or simultaneously to models.

### Sensitivity analyses

RI-CLPM estimates were generally similar when stratifying by gender (Supplementary Figure S3). Exceptions were that depressive symptoms and lower happiness were longitudinally related to higher compliance in females, with no clear association among males (though confidence intervals were wide). There was also an association between compliance and greater loneliness, depression and lower happiness at next interview among females.

Our main RI-CLPM models included linear time trends to account for secular trends in several of the studied factors (Supplementary Figure S1). We attempted more complex adjustments for time, including cubic times trends and date fixed effects, but models did not converge. Consequently, as an alternative sensitivity analysis, we instead ran fixed effects models using different adjustments for time trends. Not accounting for time was found to generate large biases in results, but estimates were very similar regardless of whether time was accounted for with linear trends, cubic trends or date fixed effects (Supplementary Figure S5).

## Discussion

This study explored predictors of compliance during the COVID-19 pandemic in a longitudinal sample of adults in the UK during the first three months of social distancing measures. Whilst there were a number of cross-sectional associations between studied factors and compliance, when looking at within-person changes, factors relating to mental health, confidence in the health service or access to essentials, social experiences, and awareness of COVID-19 were not related to future changes in compliance. The only factor that was associated future compliance was confidence in government. This suggests that cross-sectional designs – the predominant approach used in the literature – could provide substantially biased results and highlights the importance of longitudinal data in investigating predictors of compliance during pandemics. When looking at how compliance affected other factors such as mental health, there was weak evidence that compliance was related to small later increases in depressive and anxiety symptoms, predominantly among women. But we did find evidence for parallel changes in compliance and behaviours, with individuals reporting low compliance also reporting spending more time working outside the house, participating in community groups or caring for an adult, and individuals reporting high compliance also reporting more remote working.

Our finding that trust in government predicted compliance echoes findings from previous studies ^7,10– 14^. While the estimated effects are small, changes in compliance did not dissipate immediately. Further, at a population level, the consequences of small changes in compliance could be considerable, particularly if there is “social contagion” in compliance levels. This suggest that maintaining confidence is imperative for governments during pandemics. Previous work has shown that if governments make decisions that are deemed as unsafe by the public (such as easing lockdown measures too early) or if they do not uphold the same rules for government staff as for members of the public, these events can be followed by sharp declines in public confidence in government ^4^. This echoes literature showing that individuals may be less willing to comply with rules if they see others violating them, both for reciprocity reasons – a large body of evidence shows that individuals are conditional co-operators ^44^ – and as a form of social learning ^45^.

It is notable that we find little clear evidence of an association between mental health and wellbeing and compliance. This is at odds with some previous studies, which have suggested associations, albeit often in opposite directions ^17,24–27^. One possibility is that anxiety, depression and wellbeing have multiple, countervailing effects on compliance behaviours, meaning that the net association is context specific. For instance, Harper et al. ^25^ found little bivariate association between depressive symptoms and compliance, but a negative correlation once COVID-19 related fears were added into regression models. Another possibility is that previous studies (many of which have use cross-sectional data) capture reverse effects, with compliance predicting worsening of mental health. We found some evidence that compliance could lead to worse mental health in our data, particularly among women, but it is encouraging that these associations are small, so such effects, if they are occurring, are unlikely to be of clinical relevance. However, it is possible that our estimates mask a wide degree of variation. A large literature shows heterogeneous responses to many major life stressors ^46^ and the costs of compliance are unlikely to be uniform ^3^. For example, experiences of adversity, including job loss and cuts in income, have been focused on more socio-economically disadvantaged groups ^47^. Therefore, future research should explore heterogeneity in both the determinants and consequences of complying with lockdown measures.

We also found little evidence that social isolation, measured as loneliness or days without contact, has an impact on compliance. This is promising given that compliance entails lower close social contact. Again, though, it may be that average effects mask considerable heterogeneity and that net associations combine multiple countervailing effects. It is also possible that more complex modelling of cross-lagged effects – for instance using higher-order lags – may generate different results. Finally, this study showed that compliance is related to time use. The evidence that working outside the house is related to lower compliance (and conversely that remote working is related to higher compliance) underscores the importance of efforts to improve or enforce guidelines in workplaces and to financially support those whose workplaces are unsafe (e.g. through furlough schemes). But it is also notable that many leisure activities were related to higher levels of compliance. Engagement in leisure may help to reduce boredom, which is a common experience during quarantines and an important barrier to compliance ^29,48^. Indeed, it is notable that among women, compliance was related to feeling one’s activities were less worthwhile and also to increased feelings of loneliness. This suggests there could be value to supporting safe socially-distanced opportunities for leisure engagement to help individuals find worthwhile, social activities to undertake during the pandemic. This may be a particular benefit to individuals who have lost employment as they may have access to fewer purposeful activities ^49^.

This study had a number of strengths. The longitudinal design allowed us to account for two possible sources of bias: confounding via time-invariant between-person characteristics and confounding through reverse causality. While many variables shared common secular time trends, it appears that we were able to account for these sufficiently in our statistical modelling. By analysing multiple variables that displayed common secular trends, we were also provided with a second test of whether common time trends explained results. Together, our results are more supportive of a causal interpretation, though as we use observational data, it is still possible that results are explained by unobserved time-varying factors. Another strength of this study was the length of follow-up. Previous studies have used much shorter time frames and typically focused on the earlier stages of the pandemic, when enforcement was stricter and compliance was higher, on average, across the population.

However, this study also had several limitations. First, we used a single generic item of self-reported compliance with COVID-19 guidelines. While the salience of the pandemic may mean individuals recall compliance well ^50^, responses may be influenced by social desirability concerns. Less compliant individuals are also likely to be less knowledgeable about COVID-19 guidelines and so may be unable to accurately judge their own non-compliance. Both of these likely bias towards finding smaller associations. Nevertheless, at a minimum, our results show that trust in government is related to compliance *intentions*. We also did not look at specific compliance behaviours, so future work could benefit from identifying what types of behaviours people are most likely to be non-compliant on. Another limitation of our study is the possibility of selection bias. We used data from a study set-up explicitly to research COVID-19. It is likely that individuals who participated in the study had a higher interest in helping tackle the pandemic than the general population at large. This interest may manifest as a higher propensity to comply with guidelines. Another issue is that government guidelines became less stringent across the study period. Participants may have been more compliant than reported if they were unaware of current guidance. It is notable that time spent seeking COVID-19 related information declined markedly through time (Supplementary Figure S1).

Nonetheless, this study still provides the largest longitudinal exploration of predictors of compliance during the COVID-19 pandemic to date, with important implications for policy makers. In particular, the results highlight the central role of trust in determining adherence to guidelines, showing that the actions of policy makers are not just of political relevance during pandemics but are also of public health relevance as they could have had wider impacts on compliance. Confidence in the central UK government to handle the pandemic effectively has fallen markedly across the pandemic, but in other countries – including Scotland – opinions of government effectiveness have increased or remained at high levels ^4^ (see Supplementary Figure 7). This highlights that increasing citizen’s trust is within governmental control. It is vital that governments work to engage with the public and communicate plans and rules effectively to improve trust and, consequently, that social distancing rules are followed as countries enter second waves.

## Methods

### Participants

We used data from the COVID-19 Social Study; a large panel study of the psychological and social experiences of over 50,000 adults (aged 18+) in the UK during the COVID-19 pandemic. The study commenced on 21 March 2020 and involves online weekly data collection for the duration of the pandemic in the UK. The study is not random and therefore is not representative of the UK population, but does contain a well-stratified sample. The sample was recruited using three primary approaches. First, snowballing was used, including promoting the study through existing networks and mailing lists (including large databases of adults who had previously consented to be involved in health research across the UK), print and digital media coverage, and social media. Second, more targeted recruitment was undertaken focusing on (i) individuals from a low-income background, (ii) individuals with no or few educational qualifications, and (iii) individuals who were unemployed. Third, the study was promoted via partnerships with third sector organisations to vulnerable groups, including adults with pre-existing mental health conditions, older adults, carers, and people experiencing domestic violence or abuse. The study was approved by the UCL Research Ethics Committee [12467/005] and all participants gave informed consent. The study protocol and user guide (which includes full details on recruitment, retention, data cleaning, weighting and sample demographics) are available at www.covidsocialstudy.org.

Lockdown began in the UK on 23 March 2020. For these analyses, we focused on participants with 2+ interviews between 01 April – 22 June (n = 54,155, observations = 395,193). Recruitment into the study was ongoing across this period. We excluded participants with missing data on key demographic data which we use to construct survey weights (n = 2,445, observations = 14,821). We used complete case analysis as there was only a small amount of item missingness in the study.

### Measures

#### Compliance with guidelines

*Compliance with guidelines* was measured weekly using a single-item measure: “Are you following the recommendations from authorities to prevent spread of Covid-19?”. The item was measured on a seven-point Likert scale, 1 = Not at all – 7 = Very much so, and analysed as a continuous variable.

#### Predictors of Compliance

##### Mental Health and Wellbeing

*Depression* during the past week was measured using the Patient Health Questionnaire (PHQ-9); a standard 9-item instrument for diagnosing depression in primary care, with 4-point responses ranging from “not at all” to “nearly every day” (range 0-27; higher scores indicate more depressive symptoms).

*Anxiety* during the past week was measured using the Generalised Anxiety Disorder assessment (GAD-7); a well-validated 7-item tool used to screen and diagnose generalised anxiety disorder in clinical practice and research, with 4-point responses ranging from “not at all” to “nearly every day” (range 0-21; higher scores indicate more symptoms of anxiety).

*Meaning in life* and *happiness* were measured with a two single item measures: “In the past week, to what extent have you felt the things you are doing in your life are worthwhile?”; “In the past week, how happy did you feel?”. Both were measured on an 11-point scale (0 = not at all; 10 = completely) and are drawn from the UK Office for National Statistics’ Annual Population Survey which is used by the UK Government to measure national subjective wellbeing ^51^.

*Sleep quality* was measured with a single item measure, “Over the past week, how has your sleep been?”. The item was measured on a five-point Likert scale: 1 = Very good, 2 = Good, 3 = Average, 4 = Not good, 5 = Very poor. We reverse code this so higher scores into better quality sleep.

*Stressors* were captured from two questions that asked participants to select which of a list of items had caused them (a) stress (however minor) in the past week, or (b) significant stress in the past week. We create an index of worries as the sum of endorsements for: “your own safety/security”, “finances”, “losing your job/unemployment”, “getting food”, and “getting medication”, “future plans”, “boredom”. The index ranges 0-7.

##### Social Experiences

*Isolation* was measured as number of days over the past week that the participant had: not left the house or garden (*Home Isolation*), had had face-to-face contact for 15 minutes or more (*Face-to-face Isolation*), and had had a phone or video call contact with someone for 15 minutes or more (*Phone Isolation*).

*Loneliness* was measured with three items: “How often do you feel that you lack companionship?”, “How often do you feel left out?”, and “How often do you feel isolated from others?”. Each was measures on a three-point scale – 1 = hardly ever, 2 = some of the time, 3 = often. We use the sum score (range 3-9). Higher scores indicate greater loneliness.

##### Confidence in Institutions

*Confidence in (devolved) government* was measured between 21 March – 18 June using a single-item measure: “How much confidence do you have in the UK government that they can handle Covid-19 well?”. Responses to both questions were scored on a 1 (“None at all”) to 7 (“Lots”) scale. Participants from Scotland, Wales and Northern Ireland were prompted to respond about their devolved government, specifically. From 18 June, participants were instead asked separate questions on confidence in central government and confidence in their devolved government, which were scored on the same seven-point scale as previously. We used responses on central government for participants from England and responses on devolved government, otherwise.

*Confidence in the health system* and *confidence in acquiring essentials* was measured with two questions: “How much confidence do you have that the UK health service can cope during Covid-19?”; and “How much confidence do you have that essentials (e.g. access to food, water, medicines, deliveries) will be maintained during Covid-19?”. Participants from Scotland, Wales and Northern Ireland were asked to answer about the health service in their home nation, specifically. Responses to both questions were scored on a 1 (“None at all”) to 7 (“Lots”) scale.

##### COVID-19 Awareness

*Knowledge* was measured with a single item question, “How would you rate your knowledge level on Covid-19?”. Response were scored on a seven-point scale (1 = very poor knowledge, 7 = very good knowledge). *Information seeking* was measured with two items on time spent reading, watching the news, or listening to radio broadcasts about COVID-19 and time spend tweeting, blogging, or posting content online about COVID-19. Both were measured on a five-point scale (0 = did not do; 1 = <30 minutes; 2 = 30 minutes – 2 hours; 3 = 3-5 hours; 4 = 6+ hours). We used the average of responses to the two questions.

#### Time Use

Each week, participants were asked how much time they had spent in each of a list of activities during the weekday prior to interview. Time spent on each activity was collected using a five-point scale (0 = did not do; 1 = <30 minutes; 2 = 30 minutes – 2 hours; 3 = 3-5 hours; 4 = 6+ hours). To measure time spent in work, we included separate measures for time spent *remote working, working outside house, caring for friends or relatives, childcare, volunteer work* and *household chores*. To measure time spent in leisure activities, we included separate measures for *community group engagement, arts and crafts, gardening, and non-productive leisure*. The specific questionnaire items used to construct these measures are displayed in Table 2. Where we used multiple items to measure a single activity, we used the average score across items.

**Table 2:** Questionnaire items on time use

| Activity | Questionnaire Items |
| --- | --- |
| *Remote working* | - Phoning or video talking with colleagues whilst working remotely<br>- Undertaking other work remotely |
| *Working outside house* | - Going to work outside of the house (e.g. to the office) |
| *Caring for friends or relatives* | - Caring for a friend or relative |
| *Childcare* | - Caring for children (e.g. bathing, feeding, doing homework with etc)<br>- Playing with children (e.g. general play or board games or card games) |
| *Volunteer work* | - Volunteering |
| *Household chores* | - Household chores (cooking, cleaning, ironing, tidying, online shopping etc) |
| *Non-productive leisure* | - Playing video or computer games alone, or with adults or children<br>- Watching TV, films, Netflix etc (NOT for information on Covid-19)<br>- Browsing the internet (NOT for information on Covid-19)<br>- Procrastinating or not doing anything in particular |
| *Community group engagement* | - Going out of the house to engage in a community group |
| *Arts and crafts* | - Engaging in a home-based arts or crafts activity (e.g. painting, creative writing, sewing, playing music, etc)<br>- Engaging in a digital arts activity (e.g. streaming a concert, virtual tour of a museum etc)<br>- Doing DIY, woodwork, metal work, model making or similar |

### Analysis

We analysed time-use and non-time-use measures separately. To analyse the longitudinal relationship between the non-time-use measures and compliance, we estimated Random Intercept-Cross Lagged Panel Models (RI-CLPM) ^42^. The RI-CLPM is specified in a structural equation modelling framework. It adds correlated latent random intercept factors to the standard cross lagged panel model. Lagged paths are modelled between observation-level residuals once person-specific means are accounted for (see Figure 3). This allows for the separation of within-person variation from between-person variation when estimating cross-lagged effects. Accordingly, it can be used to answer the question whether *changes* in one variable are followed by changes in the other (and vice versa), rather than confounding longitudinal with cross-sectional variation, which the standard CLPM does ^42^.

**Figure 3:**
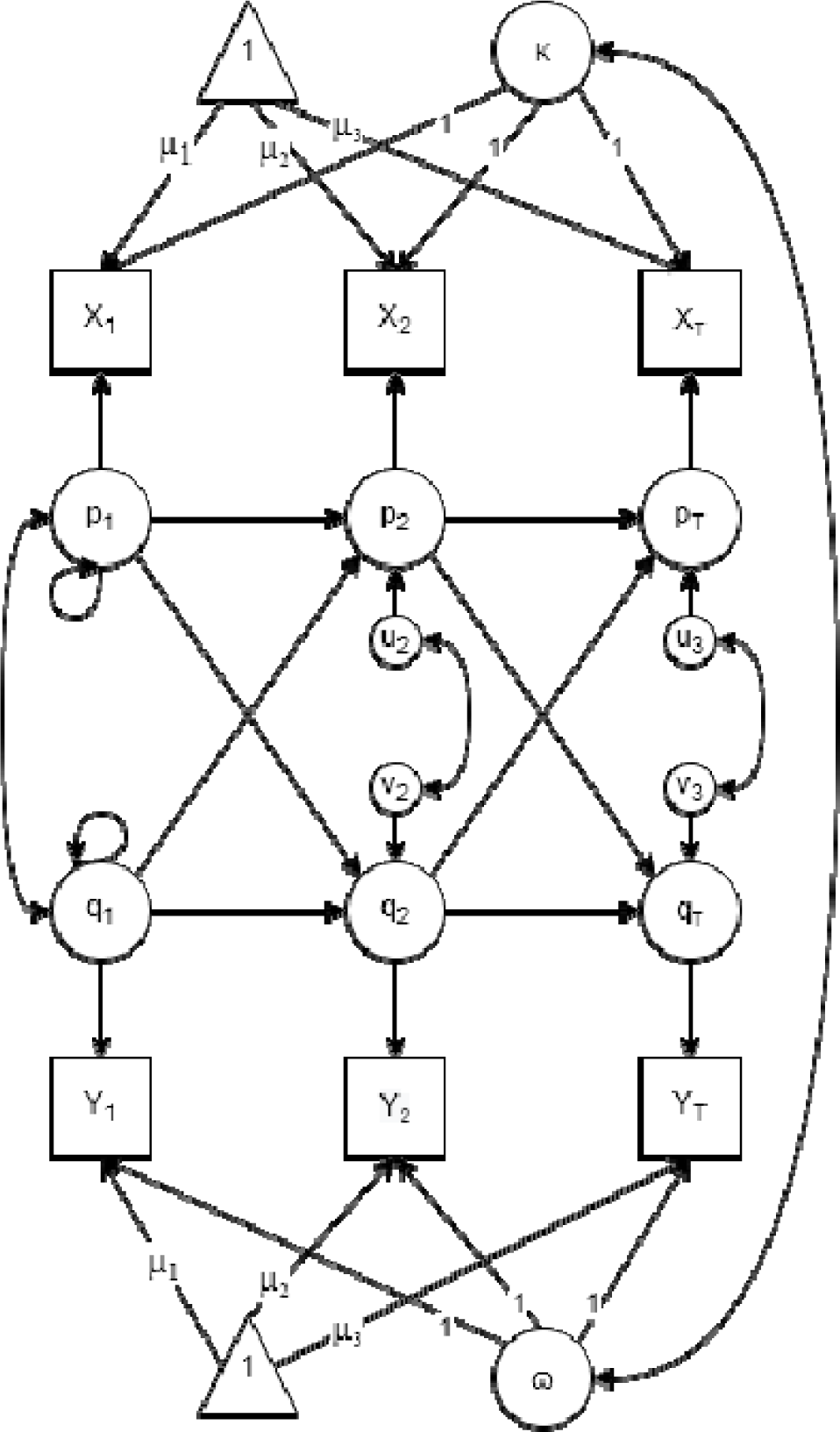
Hamaker et al. ^42^ RI-CLPM model (with means) for three-period case. Latent factors, κ and □, are random intercepts. p_t_ and q_t_ are residuals in observed values, x_t_ and y_t_, once time- and person-specific means are removed. Autoregressive and cross-lagged effects are modelled between these residuals.

We estimated a separate RI-CLPM for each non-time-use measure defined above, using the full sample and also stratifying by gender as prior work shows some differences in compliance between males and females ^28^. As individuals vary according to number of follow-ups, we used the full-information maximum likelihood (FIML) estimator in order to use data from all participants, rather than from a balanced panel. We restricted path coefficients to be equal across time, given panel imbalance and participants being able to enter the survey on different dates. We included linear time trends in the model to account for strong time trends in some of the variables in this analysis (Supplementary Information Figure S1). Our models used data from up to 11 interviews for each participant. Further details on the RI-CLPM methods is in the Supplementary Material.

To analyse the association between the time-use measures and compliance, we ran standard fixed effects regression models. These models included time use and compliance measures from the same interview, rather than lagged effects. We estimated models for each time-use measure separately and a further model including all time-use measures together. To account for time trends in the data, we added date fixed effects into each model.

RI-CLPM models were fit using the lavaan R package version 0.6-6 ^52^. As we analysed multiple variables in this study, we report Bonferroni corrected confidence intervals and standardized effect sizes (standardized using within-person standard deviation). ^1^ We included survey weights in models to account for the non-representativeness of the sample. More detail on the design of the survey weights can be found in the Supplementary Information.

Due to restrictions placed by the ethics committee, data from the COVID-19 Social Study will be made available at the end of the pandemic. The code used in this analysis is available at https://osf.io/7y9pw/.

## Supplementary Information

### Methods

Many of the variables in this analysis exhibit strong time trends (Figure S1). These trends may lead to spurious associations between compliance and the analysed factors. To account for this, we include linear time trends in the RI-CLPM models and date fixed effects in the fixed effects models. We attempted more complex adjustments for time in the RI-CLPM models, including cubic times trends and date fixed effects, but models did not converge. Therefore, as a sensitivity analysis, we instead ran fixed effects models (“within” estimator) using different adjustments for time trends as an indirect test of whether RI-CLPM results were biased due to potentially simplistic modelling of time (Figure S4). Note, the RI-CLPM model results are preferable to the fixed effects results as RI-CLPM models allow for modelling the endogeneity of the explanatory variables. The time period differs slightly between follow-ups, but is generally between 7-9 days.

As the COVID-19 Social Study used a nonprobability sampling design, we weighted data using cross-sectional weights. The weights were created using the Stata user written command ‘ebalance’. The weighted data were matched to population statistics across the following characteristics: age, gender, ethnicity, education, and country of living. Population statistics were taken from the ONS’s Annual Population Survey: (https://www.ons.gov.uk/peoplepopulationandcommunity/populationandmigration/populationestimates/datasets/populationestimatesforukenglandandwalesscotlandandnorthernireland).

We used an unbalanced panel in the models. An issue with using an unbalanced panel is that participants who remained in the survey longer contribute more data. Non-random attrition from the study may create biases, though this is partly offset by our use of within-variation when estimating cross-lagged effects. In Supplementary Figure S2, we assess attrition from the survey in detail. We ran linear OLS regression models to investigate baseline characteristics of participants who dropped out the survey by the end of the analysis period. We also ran fixed effects models to investigate within-person changes in participant characteristics (e.g. mental health) in the last interview completed before attrition from the survey. Individuals who drop out from the study comply with government guidelines less, experience more stressors, have lower confidence in government and have worse mental health and lower subjective wellbeing. However, in their last interviews’, participants report more confidence in government and better mental health and subjective wellbeing than their (participant-specific) average level. Lower compliance with guidelines is related to dropping out in both OLS and fixed effects regressions.

---

**Table S1:**
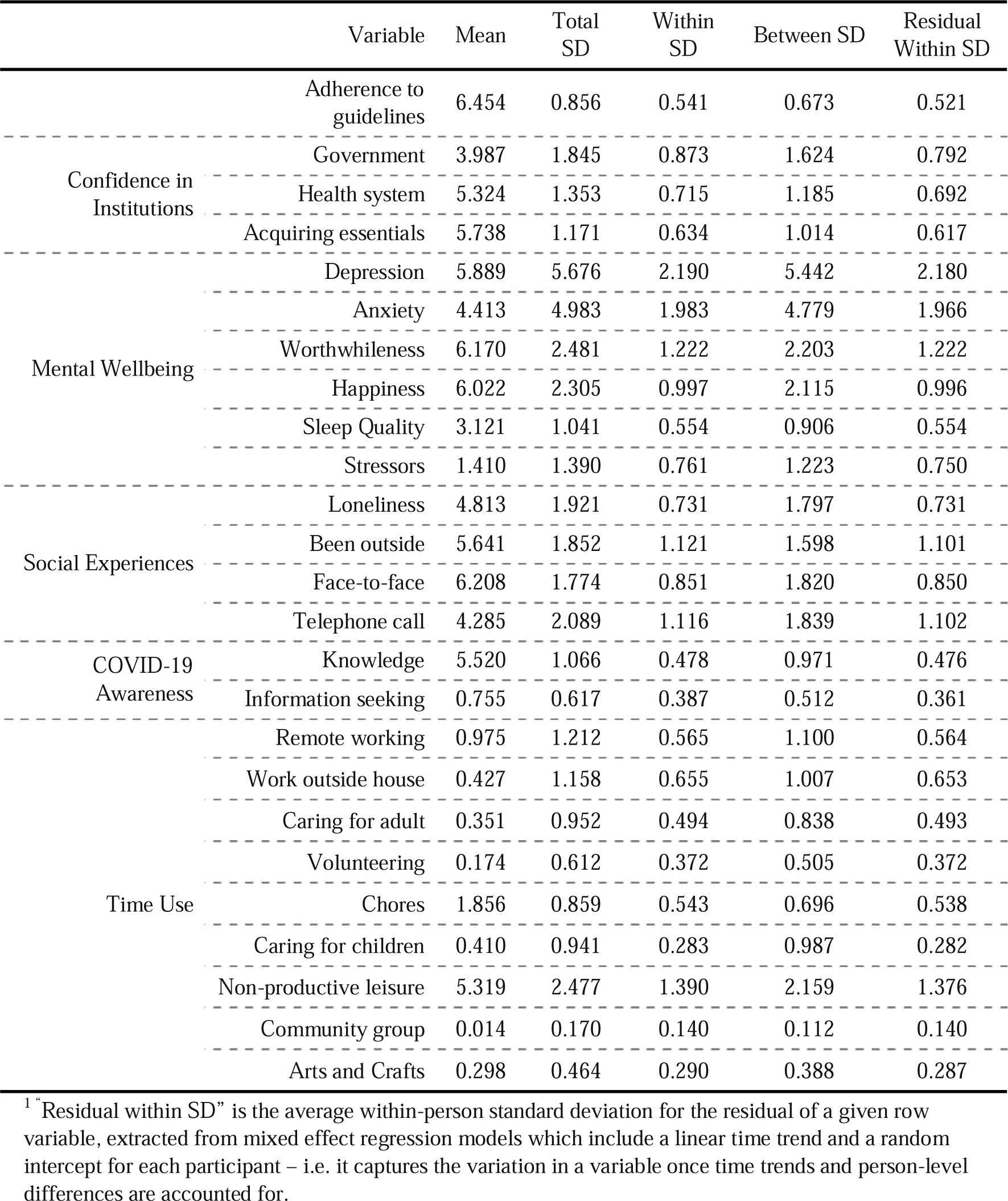
(Unweighted) descriptive statistics.

|  | Variable | Mean | Total SD | Within SD | Between SD | Residual Within SD |
| --- | --- | --- | --- | --- | --- | --- |
|  | Adherence to guidelines | 6.454 | 0.856 | 0.541 | 0.673 | 0.521 |
| Confidence in Institutions | Government | 3.987 | 1.845 | 0.873 | 1.624 | 0.792 |
|  | Health system | 5.324 | 1.353 | 0.715 | 1.185 | 0.692 |
|  | Acquiring essentials | 5.738 | 1.171 | 0.634 | 1.014 | 0.617 |
| Mental Wellbeing | Depression | 5.889 | 5.676 | 2.190 | 5.442 | 2.180 |
|  | Anxiety | 4.413 | 4.983 | 1.983 | 4.779 | 1.966 |
|  | Worthwhileness | 6.170 | 2.481 | 1.222 | 2.203 | 1.222 |
|  | Happiness | 6.022 | 2.305 | 0.997 | 2.115 | 0.996 |
|  | Sleep Quality | 3.121 | 1.041 | 0.554 | 0.906 | 0.554 |
|  | Stressors | 1.410 | 1.390 | 0.761 | 1.223 | 0.750 |
| Social Experiences | Loneliness | 4.813 | 1.921 | 0.731 | 1.797 | 0.731 |
|  | Been outside | 5.641 | 1.852 | 1.121 | 1.598 | 1.101 |
|  | Face-to-face | 6.208 | 1.774 | 0.851 | 1.820 | 0.850 |
|  | Telephone call | 4.285 | 2.089 | 1.116 | 1.839 | 1.102 |
| COVID-19 Awareness | Knowledge | 5.520 | 1.066 | 0.478 | 0.971 | 0.476 |
|  | Information seeking | 0.755 | 0.617 | 0.387 | 0.512 | 0.361 |
| Time Use | Remote working | 0.975 | 1.212 | 0.565 | 1.100 | 0.564 |
|  | Work outside house | 0.427 | 1.158 | 0.655 | 1.007 | 0.653 |
|  | Caring for adult | 0.351 | 0.952 | 0.494 | 0.838 | 0.493 |
|  | Volunteering | 0.174 | 0.612 | 0.372 | 0.505 | 0.372 |
|  | Chores | 1.856 | 0.859 | 0.543 | 0.696 | 0.538 |
|  | Caring for children | 0.410 | 0.941 | 0.283 | 0.987 | 0.282 |
|  | Non-productive leisure | 5.319 | 2.477 | 1.390 | 2.159 | 1.376 |
|  | Community group | 0.014 | 0.170 | 0.140 | 0.112 | 0.140 |
|  | Arts and Crafts | 0.298 | 0.464 | 0.290 | 0.388 | 0.287 |

<sup>1</sup>“Residual within SD” is the average within-person standard deviation for the residual of a given row variable, extracted from mixed effect regression models which include a linear time trend and a random intercept for each participant – i.e. it captures the variation in a variable once time trends and person-level differences are accounted for.

**Table S2:**
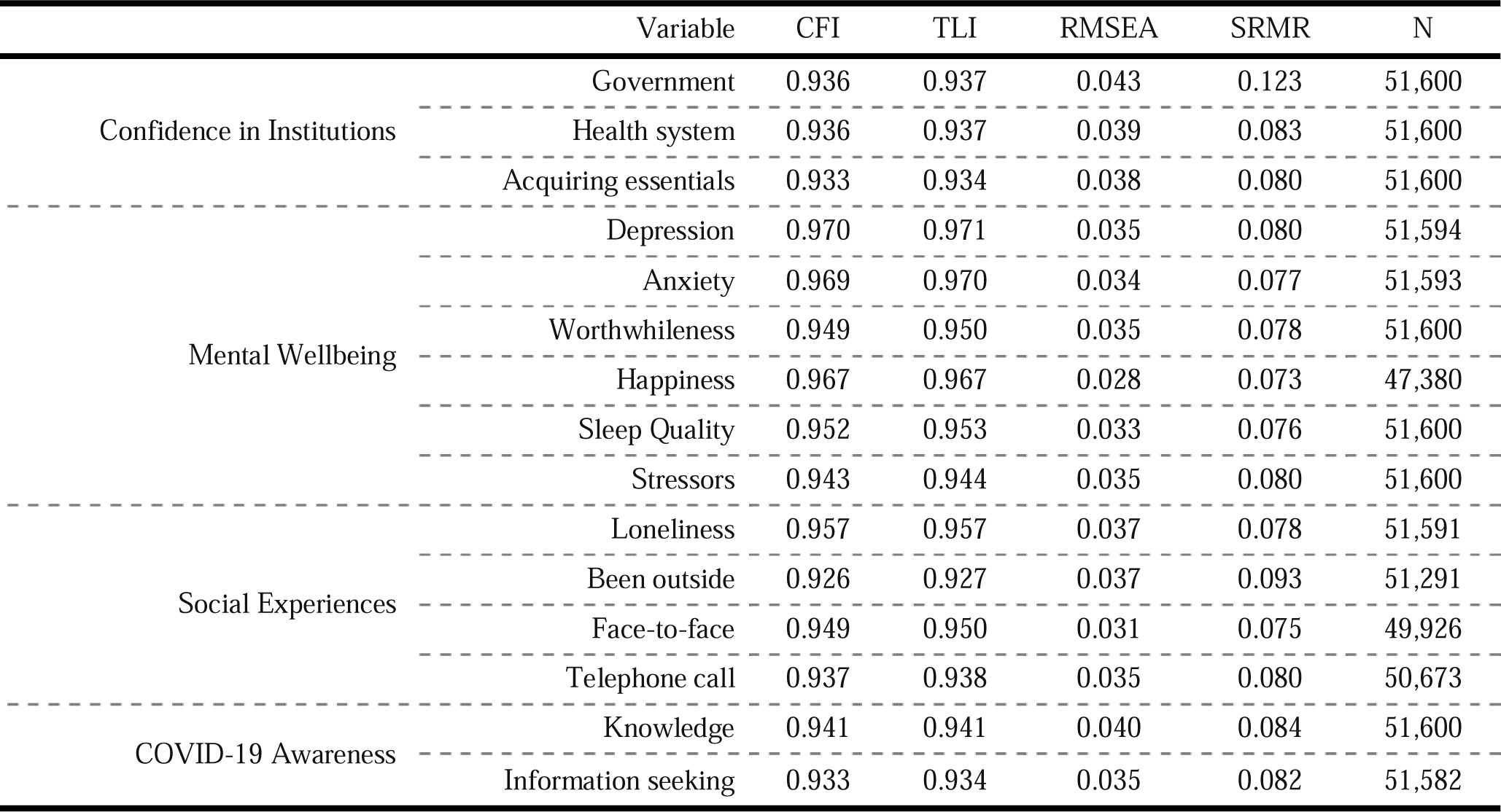
Fit Statistics, RI-CLPM models.

**Figure S1:**
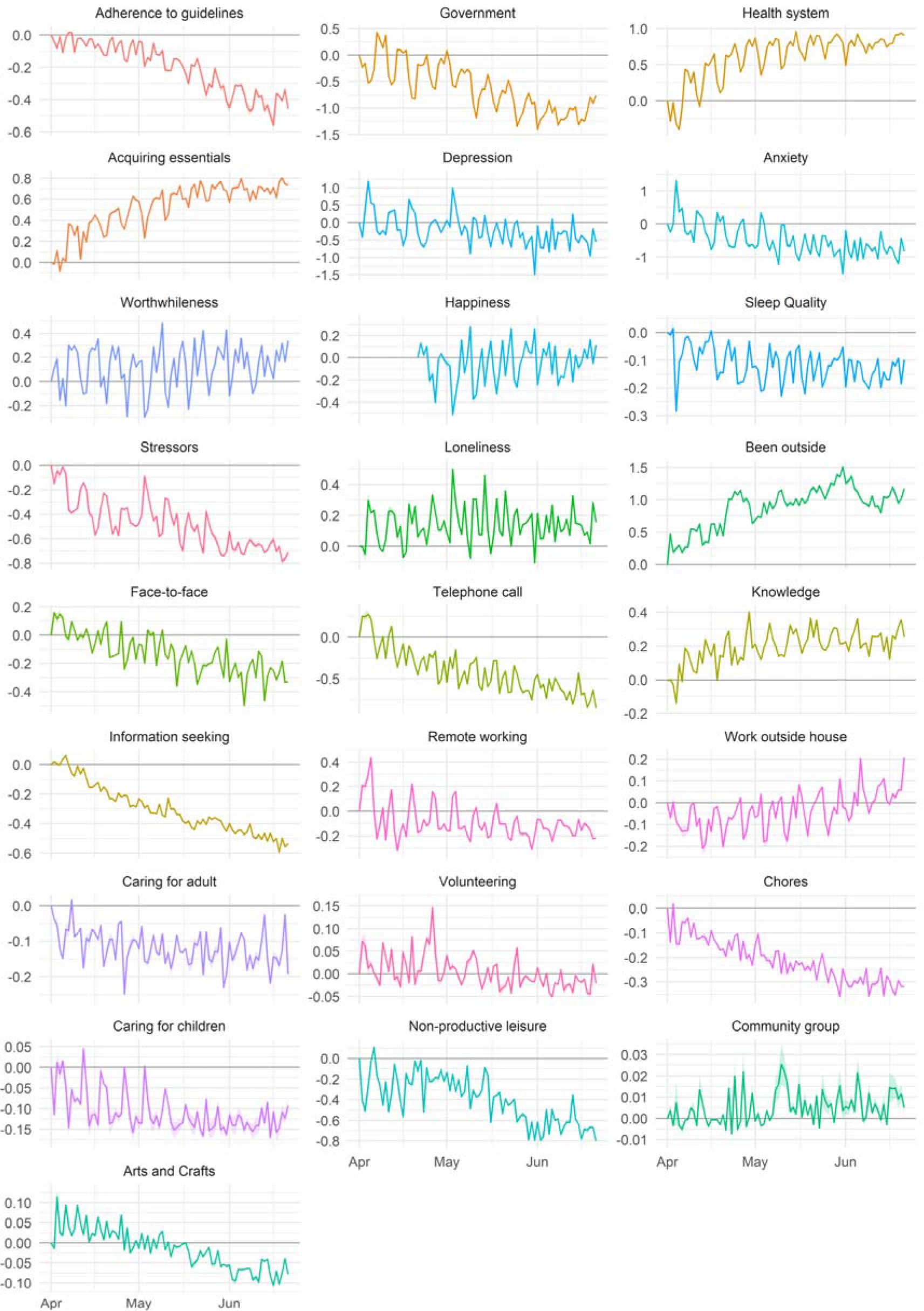
(Weighted) average daily values (+ 95% CIs) for variables used in the analysis, sample with 11 interviews between 01 April – 22 June. Scores are relative to average value on 01 April 2020.

**Figure S2:**
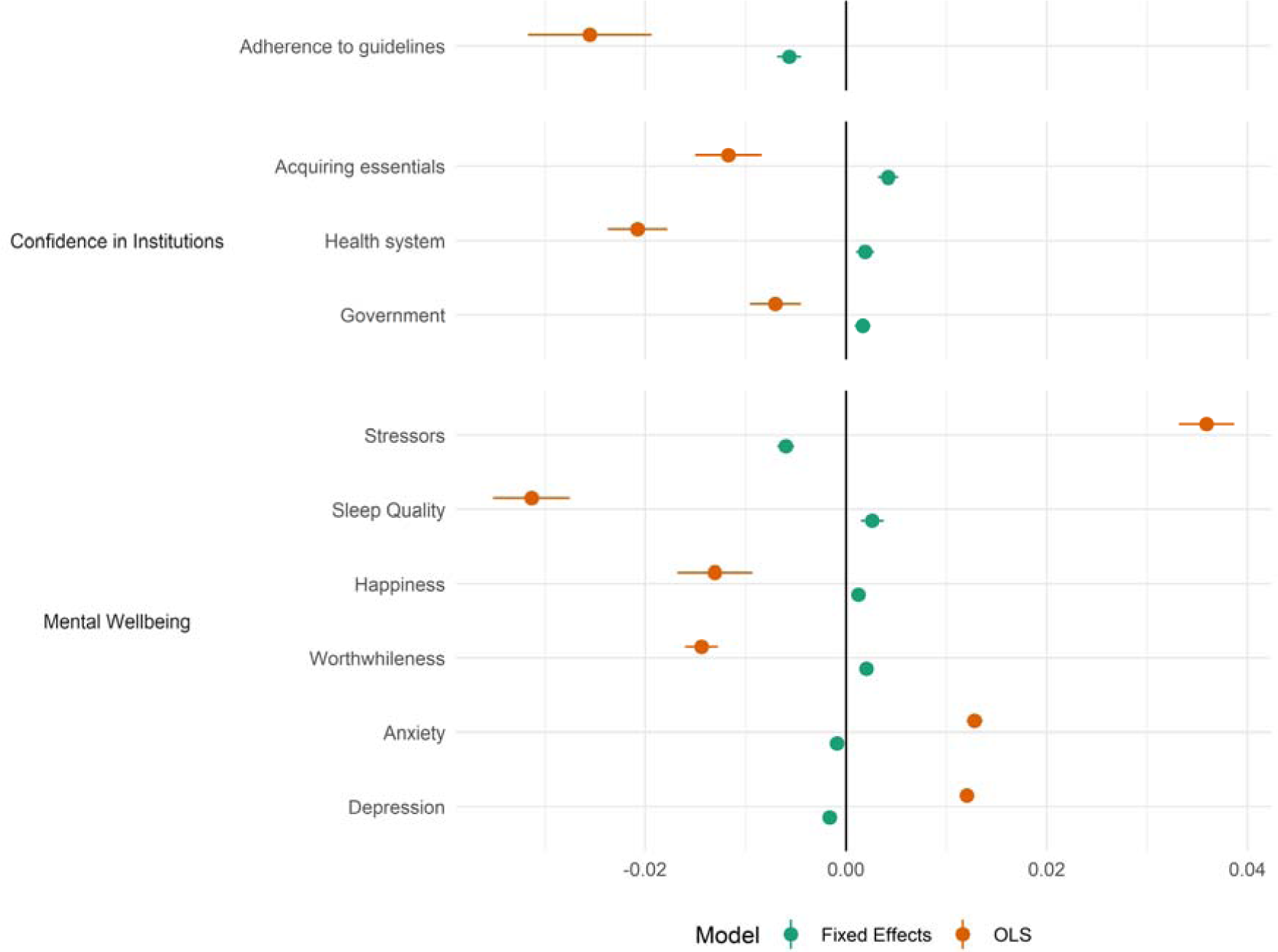
Analysis of attrition. OLS estimates refer to linear bivariate OLS regressions of attrition at any point from the survey and participant characteristics at first interview. Fixed effects estimates refer to fixed effects regression of attrition directly following interview and participant characteristics in interview. Models estimate for each characteristic separately and include cubic time trends to account for changes in drop-out through time.

**Figure S3:**
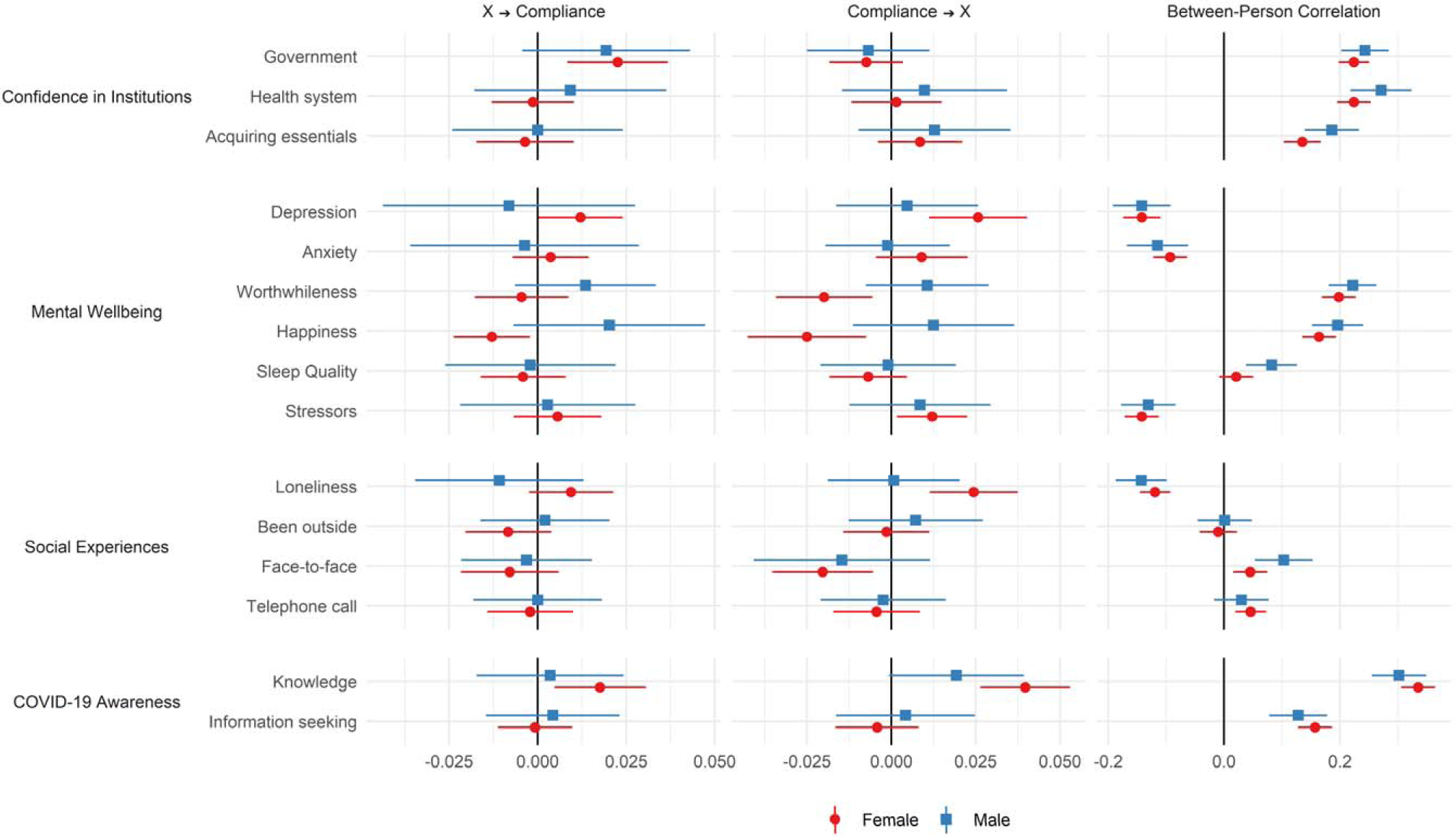
RI-CLPM Model Results stratified by gender. Left panel shows the cross-lagged effect of the exposure variable on compliance; the middle panel shows the cross-lagged of compliance on the exposure variable; and the right panel shows the correlation between the random intercept terms for the exposure and compliance

**Figure S4:**
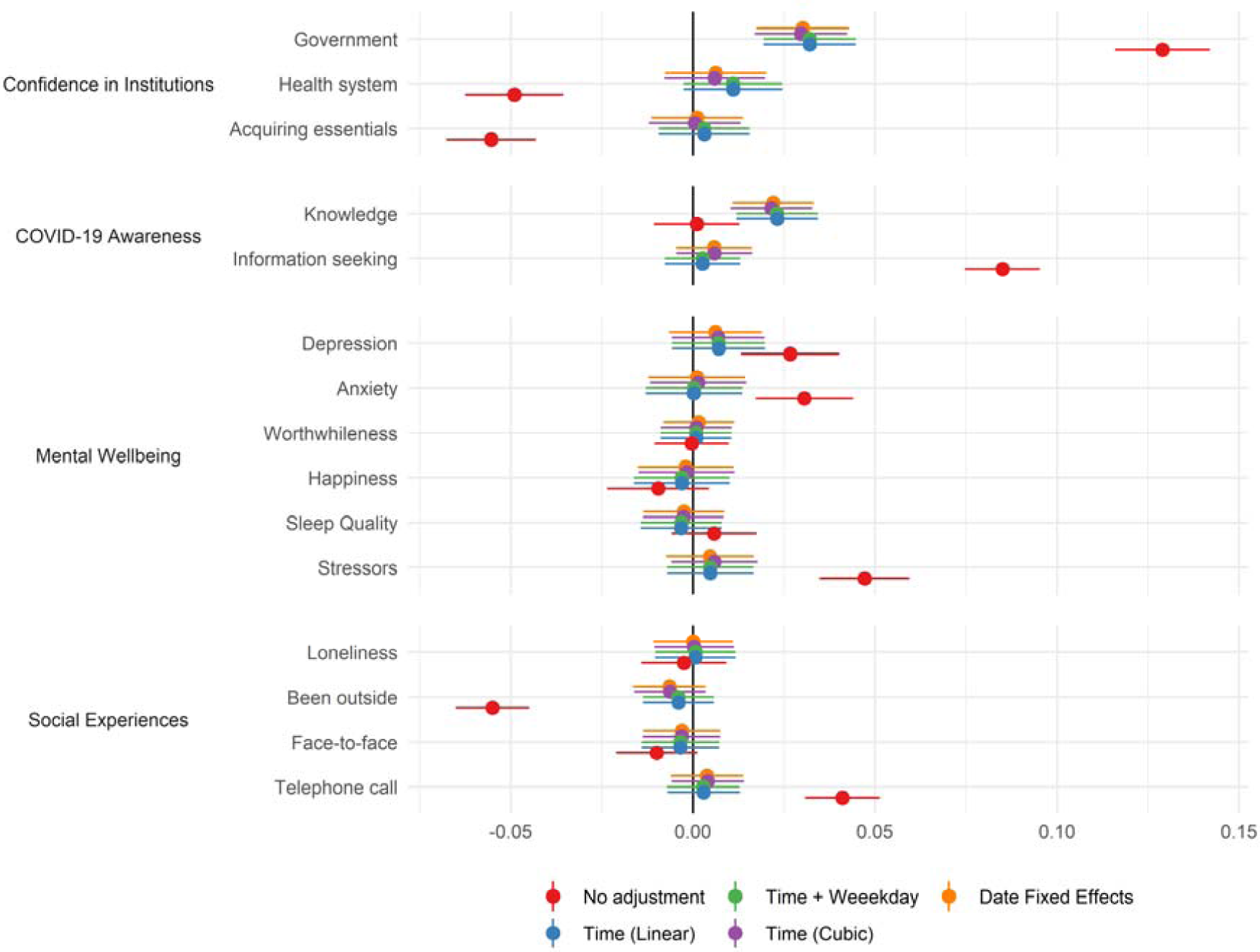
Results of fixed effects models with various methods for adjusting for time trends.

**Figure S5:**
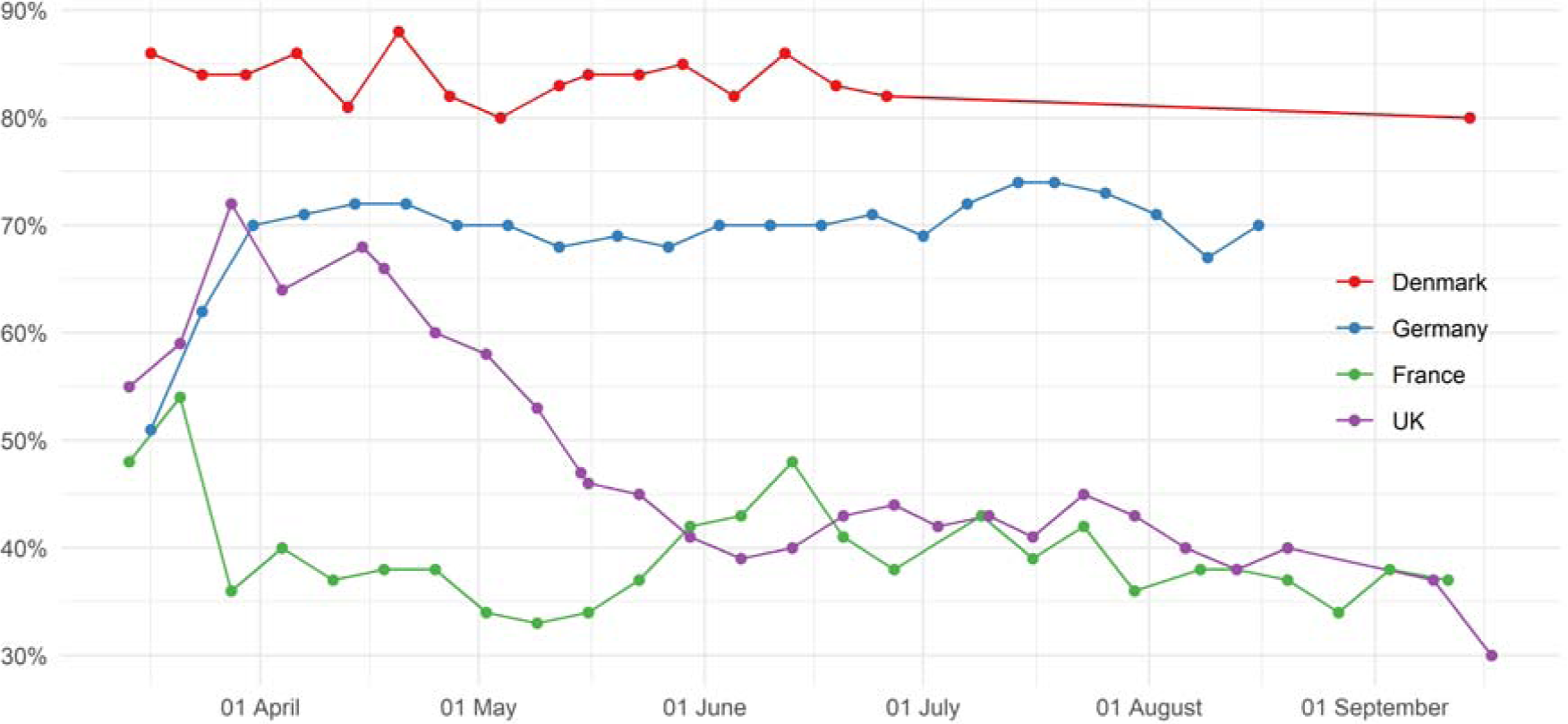
Proportion of people who think their government is handling the issue of coronavirus “very” or “somewhat” well. Source: YouGov ^53^

## Footnotes

1 We correct the non-time use measures for 15 comparisons (α = 0.05/15 = 0.0033) and the time-use measures for 9 comparisons (α = 0.05/9 = 0.0056).

